# Clinical Trial Evidence Supporting FDA Approval of Novel Therapeutic Agents Over Three Decades, 1995-2017: Cross-Sectional Analysis

**DOI:** 10.1101/19007047

**Authors:** Audrey D Zhang, Jeremy Puthumana, Nicholas S Downing, Nilay D Shah, Harlan M Krumholz, Joseph S Ross

## Abstract

**Objective:** To evaluate whether characteristics of pivotal efficacy trials supporting US Food and Drug Administration (FDA) approval of novel therapeutic agents have changed over the past three decades.

**Design:** Cross-sectional study.

**Setting and population:** Publicly available data on novel therapeutics approved by the FDA between 1995-1997, 2005-2007, and 2015-2017.

**Main outcome measures:** Use of randomization, blinding, types of comparators and primary endpoints, number of treated patients, and trial duration in pivotal trials supporting novel therapeutic approval, both individually and aggregated by each indication approval. Analyses were repeated stratifying by use of orphan designation and use of special regulatory programs.

**Results:** There were 273 novel therapeutics approved by the FDA in these 3 periods (107 in 1995-1997, 57 in 2005-2007, 109 in 2015-2017), representing 339 indications (157, 64, and 118, respectively). Overall, the proportion of indication approvals supported by at least 2 pivotal trials decreased (80.6% in 1995-1997, 60.3% in 2005-2007, 52.8% in 2015-2017; p<0.001). The proportion supported by only single-arm pivotal trials increased (4.0% in 1995-1997, 12.7% in 2005-2007, 17.0% in 2015-2017; p=0.001), as did the proportion supported by at least one pivotal trial of 6 months’ duration (25.8% in 1995-1997, 34.9% in 2005-2007, 46.2% in 2015-2017; p=0.001). When stratified by use of special regulatory programs, pivotal trial characteristics changed over time in divergent ways, both individually and when aggregated by indication approvals.

**Conclusion:** More recent FDA approvals of novel therapeutics were based on fewer pivotal trials, with less rigorous designs but longer trial durations. These findings reinforce the importance of FDA’s strategy for requiring ongoing evaluation of therapeutic safety and efficacy after approval.

**What is already known on this topic:**

- Pivotal trial characteristics supporting US Food and Drug Administration approval of novel therapeutic agents vary widely across therapeutic characteristics.
- A growing number of approvals make use of special regulatory programs that expedite the development and review of potentially transformative drugs, such as Fast Track, Priority Review, Accelerated Approval, and Breakthrough Designation.
- Special regulatory programs often offer more flexible requirements for approval, such as requiring only a single pivotal trial or trials using surrogate endpoints.

**What this study adds:**

- Over the past three decades, the characteristics of the aggregate pivotal trials supporting new drug approvals has changed, with more recent approvals requiring fewer trials of less rigorous design.
- Differences in pivotal trial characteristics over time persist after accounting for the increasing use of special regulatory programs.
- Divergent patterns are observed among approvals making use of special regulatory programs, for which more recent approvals have required fewer trials of less rigorous design, as compared to approvals using the standard pathway, for which more recent approvals have required trials of more rigorous design.

## INTRODUCTION

In the United States, the Food and Drug Administration (FDA) issues approvals for new drugs and biologics that have demonstrated safety and efficacy in “adequate and well-controlled studies”[1]. Pivotal trials are the most critical of these trials, often identified directly by FDA reviewers as the basis for approval and described in detail in FDA approval packages [1]. Early guidance suggested that at least two such trials were required for approval [2], but the FDA has maintained a flexible interpretation, taking into consideration the ethical acceptability of conducting additional trials or the rarity of diseases when determining the sufficient threshold for safety and efficacy [3]. As a result, the quantity and quality of evidence supporting recent drug approvals is variable, both in terms of the number of pivotal trials and their design features, such as randomization, blinding, choice of comparators and endpoints, number of treated patients, and trial duration [4–6].

Potentially contributing to this variability is the increasing number of special regulatory programs available to the FDA over the past 30 years, now including Fast Track (1988, in statute 1997), Priority Review (1992), Accelerated Approval (1992), and Breakthrough Designation (2012). Many of these programs codify special evidentiary standards acceptable for FDA approval of certain drugs and biologics, with the goal of promoting earlier market availability of certain therapies, such as those addressing an unmet need, or those treating serious or life-threatening conditions (**Box S1)**. As these new programs are conformed to the regulatory environment in addition to existing programs such as Orphan Designation (1983) for rare diseases [7], it is critical to understand their potential influence on the quality of evidence supporting the new drugs and biologics that clinicians prescribe to their patients.

To address this question, we examined the clinical evidence supporting FDA approval of new drugs and biologics in the following three-year periods, selected to illustrate the step-wise statutory implementation of the special regulatory programs across three decades: 1995-1997, 2005-2007, and 2015-2017. We characterized all new drug and biologic approvals within each given year, as well as the pivotal trials supporting these approvals, and determined whether trial design features differed by time period, including use of randomization, blinding, types of comparators, types of primary endpoints, number of treated patients, and trial duration. These findings will offer important insights into the influence of special regulatory programs on FDA’s evidentiary standards for new drugs and biologics over time, as well as helping patients and clinicians better understand whether the clinical evidence supporting FDA approvals has changed.

## METHODS

### Sample Construction

The FDA lists all new drug applications and biological licensing applications on the Drugs@FDA database [8]. Using a previously-described method [5], we identified new drugs and biologics (e.g. new molecular entities (NMEs) or new biologic drugs) approved between the years of 1995-1997, 2005-2007, and 2015-2017, excluding new formulations, generics, and non-therapeutic agents (e.g. diagnostic and contrast agents). We obtained the complete action package for the original approval of each therapeutic, either through the Drugs@FDA database, or in the case of the 1995-1997 therapeutics, through a Freedom of Information Act (FOIA) request.

### Therapeutic Characteristics

Based on information from the approval package, we classified each novel therapeutic by period of approval and product type (small-molecule or biologic). Using information available in the approval packages and from public listings available on the FDA website, we identified whether each therapeutic was evaluated through a special regulatory program (Priority Review, Accelerated Approval, Fast Track, Breakthrough Therapy) [9–12]. These special regulatory programs are used for therapeutics that are intended to address “unmet medical needs” for serious or life-threatening conditions [13]. Data on certain special regulatory programs was not available in all years; data for Fast Track was only available after 1997, when it was codified by the 1997 Food and Drug Administration Modernization Act (FDAMA), and Breakthrough Therapy was only introduced in 2012 by the Food and Drug Administration Safety and Innovation Act (FDASIA) [14].

Using information available in the approval packages, we identified the indications for each novel therapeutic at the time of initial approval. We classified these into one of eight therapeutic areas based on the World Health Organization’s Anatomic Therapeutic Classification System [15]. Using the Orphan Products Designation Database, we also determined whether these originally-approved indications had been granted orphan status, a designation granted at sponsor request to drugs for indications for which there are 200,000 or fewer patients in the United States, indications for which alternative therapeutic options are often not available [16].

### Trial Characteristics

We followed a previously-described method to identify and characterize the pivotal efficacy trials used as the basis of approval for each indication of each new drug or biologic [4,5]. Briefly, these were generally labeled as “pivotal” by FDA medical reviewers; in cases where they were not explicitly labeled, we identified these trials using FDA medical reviewer descriptions of clinical studies, including those trials described as essential to approval or those highlighted individually for discussion of study design or analysis of study results. Additionally, any new efficacy trial reviewed as part of a resubmitted application was considered pivotal to approval.

For each identified trial, we determined its use of randomization and blinding, categorized as randomized vs. non-randomized and double-blinded vs. not double-blinded, respectively. Next, we categorized use of a comparator as active treatment, placebo control, or none, in the case of single-arm trials. We categorized primary trial endpoints as clinical endpoints, clinical scales, or surrogate endpoints using a previously developed framework [5,17]. Briefly, clinical endpoints, such as death or hospitalization, are those that measure patient-reported outcomes, function, or survival; clinical scales, such as the visual analog scale for pain, represent ordinal characterization of symptoms; and surrogate endpoints, such as hemoglobin A1c, represent biomarkers expected to predict clinical benefit. We determined the number of treated patients by abstracting the number of patients included in intention-to-treat (ITT) analyses. We also determined the duration of each trial. For time-driven endpoints, duration was defined as the time of primary endpoint measurement, such as hemoglobin A1c at 24 weeks. For event-driven endpoints, such as progression-free survival, duration was defined as the median follow-up time for participants, or a weighted average of the median follow-up time in cases where it differed between trial arms. Initial abstraction was performed by 3 investigators (ADZ, JP, NSD).

### Statistical Analysis

We used descriptive statistics to characterize the sample of new drugs and biologics and their indications for use, and to characterize the features of their supporting pivotal trials. We used chi-squared tests for trend and Spearman rank tests as appropriate to compare trial characteristics across the three periods of interest, first at the level of individual pivotal trials and then considering the aggregate pivotal trial evidence supporting each indication approval. Analyses aggregated by indication included only those indications for which pivotal trial characteristic reporting was complete, to prevent undercounting in cases with missing data. We stratified analyses by use of any special regulatory program, as defined above, and by use of orphan designation.

As supplementary analyses, we repeated analyses stratified by product type (small-molecule vs. biologic) and therapeutic area. We conducted sensitivity analyses considering the effect of expected length of treatment on duration. Expected length of treatment was determined categorized as acute for those whose expected length of use was less than 1 month, intermediate for those between 1 month and 2 years, and chronic for those greater than 2 years. We also conducted sensitivity analyses examining the individual regulatory programs spanning three time periods (e.g. Priority Review and Accelerated Approval). All statistical tests were 2-tailed and used the Bonferroni method to adjust p-values to account for multiple comparisons. All analyses were conducted using R version 3.5.1 (R Foundation for Statistical Computing).

### Patient Involvement

No patients were involved in setting the research question or the outcome measures, nor were they involved in developing the design or implementation of the study. Our findings will be made available to patients, but there are no plans to partner with patients for dissemination. Because this study did not use patient data, it was exempt from review by the Yale Human Investigation Committee.

## RESULTS

### Study Sample

We identified 273 new drugs and biologics approved by the FDA for 339 indications across three periods: 107 drugs for 157 indications in 1995-1997, 57 drugs for 64 indications in 2005-2007, and 109 drugs for 118 indications in 2015-2017. Product and indication characteristics differed across these periods **(Table 1)**. The proportion of biologics represented among new approvals has increased, as have the proportion of approvals using any special regulatory program (34.6% in 1995-1997, 57.9% in 2005-2007, 64.2% in 2015-2017) or orphan designation (12.7% in 1995-1997, 26.6% in 2005-2007, 38.1% in 2015-2017; all p<0.001). The therapeutic areas associated with new indication approvals have shifted, with the most common therapeutic area being infectious disease in 1995-1997 (n=53, 33.8%), oncology in 2015-2017 (n=32, 27.1%) **(Table 1)**.

**Table 1.** Characteristics of New Drugs and Biologics Approved by the U.S. Food and Drug Administration in 1995-1997, 2005-2007, and 2015-2017.

|  | **1995-1997** | **2005-2007** | **2015-2017** |  |
| --- | --- | --- | --- | --- |
| **Product Characteristics** | **107** | **57** | **109** |  |
| *# Indications, Median (IQR, Range)* | 1 (1-1, 1-14) | 1 (1-1, 1-6) | 1 (1-1, 1-2) | P=0.028 |
| *Agent Type, No. (%)* |  |  |  |  |
| Drugs | 104 (97.2) | 49 (86.0) | 79 (72.5) | P<0.001 |
| Biologics | 3 (2.8) | 8 (14.0) | 30 (27.5) | P<0.001 |
| *Special Regulatory Program, No. (%)* |  |  |  |  |
| Any | 37 (34.6) | 33 (57.9) | 70 (64.2) | P<0.001 |
| Priority Review | 36 (33.6) | 31 (54.4) | 67 (61.5) | P<0.001 |
| Accelerated Approval | 12 (11.2) | 12 (21.1) | 18 (16.5) | P=0.28 |
| Fast Track | - | 14 (24.6) | 39 (35.8) |  |
| Breakthrough Therapy | - | - | 34 (31.2) |  |
| **Indication Characteristics** | **157** | **64** | **118** |  |
| *Therapeutic Area, No. (%)* |  |  |  |  |
| Infectious Disease | 53 (33.8) | 16 (25.0) | 17 (14.4) | P<0.001 |
| Cancer | 17 (10.8) | 11 (17.2) | 32 (27.1) | P<0.001 |
| Cardiovascular/Diabetes/Lipids | 26 (16.6) | 10 (15.6) | 17 (14.4) | P=0.63 |
| Other | 61 (38.9) | 27 (42.2) | 52 (44.1) | P=0.38 |
| *Orphan Status, No. (%)* | 20 (12.7) | 17 (26.6) | 45 (38.1) | P<0.001 |

### Features of Individual Pivotal Trials

We identified a total of 802 pivotal trials supporting the new drugs and biologics in our sample: 408 trials in 1995-1997, 141 trials in 2005-2007, and 253 trials in 2015-2017. Of these pivotal trials, the majority were randomized **(Table 2)**, though randomization decreased from 93.6% in 1995-1997 to 82.2% in 2005-2007 and 2015-2017 (p<0.001). Likewise, most trials were double-blinded, though double-blinding decreased from 79.4% in 1995-1997 to 67.4% in 2005-2007 and 67.6% in 2015-2017 (p<0.001). Choice of comparators differed by time period, as use of active comparators decreased (44.1% in 1995-1997, 34.0% in 2005-2007, and 29.2% in 2015-2017; p<0.001) while single-arm trials increased (8.5% in 1995-1997, 17.7% in 2005-2007, and 17.8% in 2015-2017; p<0.001). Sensitivity analyses were conducted examining rates of randomization and blinding only among trials using comparators and showed no changes over time (**Table S2)**.

**Table 2.** Characteristics of Pivotal Trials Supporting New Drugs and Biologics Approved by the U.S. Food and Drug Administration in 1995-1997, 2005-2007, and 2015-2017, Stratified by Drug/Indication Characteristics.

|  | Randomiz<br>ed | Double-Blinded | Comparator |  |  | Endpoints |  |  |
| --- | --- | --- | --- | --- | --- | --- | --- | --- |
| Characteri<br>stics<br>(Pivotal<br>Trials) |  |  | Active | Placebo | None | Clinical | Scale | Surrogate |
| **Overall** |  |  |  |  |  |  |  |  |
| 1995-1997<br>(n=401) | 93.6<br>(90.7-95.8) | 79.4<br>(75.0-83.3) | 44.1<br>(39.2-49.2) | 47.4<br>(42.4-52.4) | 8.5<br>(5.9-11.6) | 43.8<br>(38.8-48.8) | 8.0<br>(5.5-11.1) | 48.3<br>(43.3-53.3) |
| 2005-2007<br>(n=141) | 82.2<br>(74.9-88.2) | 67.4<br>(58.8-75.0) | 34.0<br>(26.3-42.5) | 48.2<br>(39.7-56.8) | 17.7<br>(11.8-25.1) | 28.4<br>(21.1-36.6) | 11.3<br>(6.6-17.8) | 60.3<br>(51.7-68.4) |
| 2015-2017<br>(n=253) | 82.2<br>(76.9-86.7) | 67.6<br>(61.4-73.3) | 29.2<br>(23.7-35.3) | 53.0<br>(46.6-59.2) | 17.8<br>(13.3-23.1) | 23.3<br>(18.3-29.0) | 17.4<br>(12.9-22.6) | 59.3<br>(53.0-65.4) |
| 3-way p | P<0.001* | P<0.001* | P<0.001* | P=0.17 | P<0.001* | P<0.001* | P<0.001* | P=0.004* |
| 2-way p** | P<0.001* | P<0.001* | P<0.001* |  |  | P<0.001* |  |  |
| **Regulatory**<br>**Program** |  |  |  |  |  |  |  |  |
| Any |  |  |  |  |  |  |  |  |
| 1995-1997<br>(n=89) | 80.9<br>(71.2-88.5) | 74.4<br>(63.6-83.4) | 37.6<br>(27.4-48.8) | 43.5<br>(32.8-54.7) | 18.8<br>(11.1-28.8) | 20.0<br>(12.1-30.1) | 5.9<br>(1.9-13.2) | 75.3<br>(64.7-84.0) |
| 2005-2007<br>(n=64) | 75.0<br>(62.6-85.0) | 56.3<br>(43.3-68.6) | 35.9<br>(24.3-48.9) | 39.1<br>(27.1-52.1) | 25.0<br>(15.0-37.4) | 37.5<br>(25.7-50.5) | 7.8<br>(2.6-17.3) | 54.7<br>(41.7-67.2) |
| 2015-2017<br>(n=128) | 67.2<br>(58.3-75.2) | 53.9<br>(44.9-62.8) | 23.4<br>(16.4-31.7) | 43.8<br>(35.0-52.8) | 32.8<br>(24.8-41.7) | 19.5<br>(13.1-27.5) | 13.3<br>(7.9-20.4) | 67.2<br>(58.3-75.2) |
| 3-way p | P=0.02* | P=0.004* | P=0.02* | P=0.92 | P=0.02* | P=0.71 | P=0.07 | P=0.32 |
| 2-way p** | P=0.004* | P=0.003* | P=0.03 |  |  | P=0.22 |  |  |
| None |  |  |  |  |  |  |  |  |
| 1995-1997<br>(n=316) | 96.1<br>(93.3-98.0) | 80.7<br>(75.9-84.9) | 45.9<br>(40.3-51.6) | 48.4<br>(42.8-54.1) | 5.7<br>(3.4-8.9) | 50.2<br>(44.5-55.8) | 8.6<br>(5.7-12.2) | 41.3<br>(35.8-46.9) |
| 2005-2007<br>(n=77) | 88.3<br>(79.0-94.5) | 76.6<br>(65.6-85.5) | 32.5<br>(22.2-44.1) | 55.8<br>(44.1-67.2) | 11.7<br>(5.5-21.0) | 22.1<br>(13.4-33.0) | 14.3<br>(7.4-24.1) | 64.9<br>(53.2-75.5) |
| 2015-2017<br>(n=125) | 97.6<br>(93.1-99.5) | 81.6<br>(73.7-88.0) | 35.2<br>(26.9-44.2) | 62.4<br>(53.3-70.9) | 2.4<br>(0.5-6.9) | 27.2<br>(19.6-35.9) | 21.6<br>(14.7-29.8) | 51.2<br>(42.1-60.2) |
| 3-way p | P=0.92 | P=0.96 | P=0.02 | P=0.007* | P=0.38 | P<0.001* | P<0.001* | P=0.014 |
| 2-way p** | P=0.44 | P=0.83 | P=0.02* |  |  | P<0.001* |  |  |
| **Orphan** |  |  |  |  |  |  |  |  |
| Yes |  |  |  |  |  |  |  |  |
| 1995-1997<br>(n=36) | 80.6<br>(62.5-92.5) | 61.3<br>(42.2-78.2) | 25.8<br>(11.9-44.6) | 45.2<br>(27.3-64.0) | 29.0<br>(14.2-48.0) | 48.4<br>(30.2-66.9) | 9.7<br>(2.0-25.8) | 41.9<br>(24.5-60.9) |
| 2005-2007<br>(n=24) | 45.8<br>(25.6-67.2) | 29.2<br>(12.6-51.1) | 12.5<br>(2.7-32.4) | 33.3<br>(15.6-55.3) | 54.2<br>(32.8-74.4) | 37.5<br>(18.8-59.4) | 0.0<br>(0.0-14.2) | 62.5<br>(40.6-81.2) |
| 2015-2017<br>(n=63) | 52.4<br>(39.4-65.1) | 39.7<br>(27.6-52.8) | 6.3<br>(1.8-15.5) | 46.0<br>(33.4-59.1) | 47.6<br>(34.9-60.6) | 19.0<br>(10.2-30.9) | 7.9<br>(2.6-17.6) | 73.0<br>(60.3-83.4) |
| 3-way p | P=0.02* | P=0.09 | P=0.009* | P=0.80 | P=0.13 | P=0.003* | P=0.94 | P=0.004* |
| 2-way p** | P=0.009* | P=0.05 | P=0.02* |  |  | P=0.009* |  |  |
| No |  |  |  |  |  |  |  |  |
| 1995-1997<br>(n=372) | 94.7<br>(91.9-96.8) | 80.9<br>(76.5-84.9) | 45.7<br>(40.5-50.9) | 47.6<br>(42.4-52.8) | 6.8<br>(4.4-9.8) | 43.4<br>(38.2-48.6) | 7.9<br>(5.3-11.1) | 48.8<br>(43.6-54.0) |
| 2005-2007<br>(n=117) | 89.7<br>(82.8-94.6) | 75.2<br>(66.4-82.7) | 38.5<br>(29.6-47.9) | 51.3<br>(41.9-60.6) | 10.3<br>(5.4-17.2) | 26.5<br>(18.8-35.5) | 13.7<br>(8.0-21.3) | 59.8<br>(50.4-68.8) |
| 2015-2017<br>(n=190) | 92.1<br>(87.3-95.5) | 76.8<br>(70.2-82.6) | 36.8<br>(30.0-44.1) | 55.3<br>(47.9-62.5) | 7.9<br>(4.5-12.7) | 24.7<br>(18.8-31.5) | 20.5<br>(15.0-27.0) | 54.7<br>(47.4-62.0) |
| 3-way p | P=0.17 | P=0.21 | P=0.04 | P=0.08 | P=0.53 | P<0.001* | P<0.001* | P=0.12 |
| 2-way p** | P=0.22 | P=0.26 | P=0.14 |  |  | P<0.001* |  |  |

\* indicates $p < 0.025$ (after Bonferroni adjustment for 2 comparisons)
\*\* 2-way p-value for differences between 1995-1997 and 2015-2017 time periods.

Choice of primary endpoints also differed over time, as use of clinical endpoints decreased (43.8% in 1995-1997, 28.4% in 2005-2007, and 23.3% in 2015-2017; p<0.001) while use of surrogate endpoints increased (48.3% in 1995-1997, 60.3% in 2005-2007, and 59.3% in 2015-2017; p=0.004). Median number of treated patients in each pivotal trial also increased (277 [IQR 150-442] in 1995-1997, 404 [IQR 189-622] in 2005-2007, and 467 [IQR 209-722] in 2015-2017; p<0.001), as did median trial duration (11.0 weeks [IQR 4.9-24.0] in 1995-1997, 16.0 weeks [IQR 6.0-26.0] in 2005-2007, and 24.0 weeks [IQR 12.0-37.6] in 2015-2017; p<0.001).

### Features of Aggregated Pivotal Trials Supporting Indication Approvals

Overall, 7 (2.0%) of 339 indication approvals were not supported by any pivotal trial (**Table S3)**. Of the 332 indication approvals supported by pivotal trials, 293 (88.3%) had complete reporting of all abstracted pivotal trial characteristics within the FDA documentation (82.7% in 1995-1997, 98.4% in 2005-2007, and 89.8% in 2015-2017; **Tables S4-5)**. Among these 293 indication approvals, the proportion supported by at least 2 pivotal trials decreased over time (80.6% in 1995-1997, 60.3% in 2005-2007, and 52.8% in 2015-2017; p<0.001) (**Table 4**). The proportion of indication approvals supported only by single-arm trials increased over time (4.0% in 1995-1997, 12.7% in 2005-2007, 17.0% in 2015-2017; p=0.001). The proportion of indication approvals supported only by trials using surrogate endpoints was not statistically different over time (p=0.36), nor was the median aggregated number of treated patients (p=0.89) **(Tables 4-5)**. The proportion of indication approvals with at least one trial of 6 months’ duration increased (25.8% in 1995-1997, 34.9% in 2005-2007, 46.2% in 2015-2017; p=0.001).

**Table 3.** Number of Treated Patients and Duration of Pivotal Trials Supporting New Drugs and Biologics Approved by the U.S. Food and Drug Administration in 1995-1997, 2005-2007, and 2015-2017.

| **Characteristics**<br>**(Pivotal Trials)** | **Number of Treated Patients,**<br>**Median (IQR)** |  | **Duration in Weeks,**<br>**Median (IQR)** |
| --- | --- | --- | --- |
|  | **Overall** | **Intervention** |  |
| **Overall** |  |  |  |
| 1995-1997 (n=408) | 277 (150-442) | 168 (91-274) | 11.0 (4.9-24.0) |
| 2005-2007 (n=141) | 404 (189-622) | 243 (136-381) | 16.0 (6.0-26.0) |
| 2015-2017 (n=253) | 467 (209-722) | 279 (143-451) | 24.0 (12.0-37.6) |
| 3-way p | P<0.001* | P<0.001* | P<0.001* |
| 2-way p** | P<0.001* | P<0.001* | P<0.001* |
| **Regulatory**<br>**Program** |  |  |  |
| **Any** |  |  |  |
| 1995-1997 (n=89) | 236 (95-424) | 148 (64-259) | 24.0 (16.0-52.0) |
| 2005-2007 (n=64) | 329 (115-605) | 225 (106-336) | 17.0 (9.7-28.3) |
| 2015-2017 (n=128) | 239 (121-658) | 177 (87-366) | 24.0 (12.0-50.3) |
| 3-way p | P=0.10 | P=0.03 | P=0.61 |
| 2-way p** | P=0.07 | P=0.02* | P=0.33 |
| **None** |  |  |  |
| 1995-1997 (n=316) | 290 (165-451) | 182 (97-289) | 8.0 (4.0-16.0) |
| 2005-2007 (n=77) | 455 (273-651) | 254 (163-409) | 12.0 (5.0-26.0) |
| 2015-2017 (n=125) | 546 (427-931) | 329 (244-574) | 24.0 (12.0-26.0) |
| 3-way p | P<0.001* | P<0.001* | P<0.001* |
| 2-way p** | P<0.001* | P<0.001* | P<0.001* |
| **Orphan** |  |  |  |
| **Yes** |  |  |  |
| 1995-1997 (n=36) | 155 (51-270) | 94 (28-199) | 24.5 (7.9-78.2) |
| 2005-2007 (n=24) | 92 (55-190) | 76 (42-150) | 21.3 (8.5-52.0) |
| 2015-2017 (n=63) | 129 (69-378) | 113 (60-242) | 34.6 (14.9-70.7) |
| 3-way p | P=0.71 | P=0.20 | P=0.08 |
| 2-way p** | P=0.98 | P=0.30 | P=0.19 |
| **No** |  |  |  |
| 1995-1997 (n=372) | 290 (158-446) | 179 (97-285) | 8.1 (4.8-24.0) |
| 2005-2007 (n=117) | 471 (296-652) | 265 (173-401) | 12.0 (5.2-26.0) |
| 2015-2017 (n=190) | 543 (352-818) | 323 (220-503) | 20.0 (12.0-26.0) |
| 3-way p | P<0.001* | P<0.001* | P<0.001* |
| 2-way p** | P<0.001* | P<0.001* | P<0.001* |

\* indicates p<0.025 (after Bonferroni adjustment for 2 comparisons)
\*\* 2-way p-value for differences between 1995-1997 and 2015-2017 time periods.

**Table 4.** Characteristics of Aggregated Pivotal Trials Supporting U.S. Food and Drug Administration Indication Approvals for New Drugs and Biologics in 1995-1997, 2005-2007, and 2015-2017, Stratified by Drug/Indication Characteristics.

| Characteristics (Indications) | ≥2 Trials | Duration |  | Comparators |  | Endpoints |  |
| --- | --- | --- | --- | --- | --- | --- | --- |
|  |  | ≥1 Trial of ≥6 Months | ≥1 Trial of ≥12 Months | ≥1 Trial Using Any Comparator | Single-Arm Only | ≥1 Trial Using Clinical Endpoints | Trials Using Surrogate Endpoints Only |
| **Overall** |  |  |  |  |  |  |  |
| 1995-1997 (n=124) | 80.6<br>(72.6-87.2) | 25.8<br>(18.4-34.4) | 18.5<br>(12.1-26.5) | 96.0<br>(90.8-98.7) | 4.0<br>(1.3-9.2) | 60.5<br>(51.3-69.1) | 39.5<br>(30.9-48.7) |
| 2005-2007 (n=63) | 60.3<br>(47.2-72.4) | 34.9<br>(23.3-48.0) | 19.0<br>(10.2-30.9) | 87.3<br>(76.5-94.4) | 12.7<br>(5.6-23.5) | 49.2<br>(36.4-62.1) | 50.8<br>(37.9-63.6) |
| 2015-2017 (n=106) | 52.8<br>(42.9-62.6) | 46.2<br>(36.5-56.2) | 35.8<br>(26.8-45.7) | 83.0<br>(74.5-89.6) | 17.0<br>(10.4-25.5) | 54.7<br>(44.8-64.4) | 45.3<br>(35.6-55.2) |
| 3-way p | P<0.001* | P=0.001* | P=0.003* | P=0.001* |  | P=0.36 |  |
| 2-way p** | P<0.001* | P=0.001* | P=0.003* | P=0.001* |  | P=0.38 |  |
| **Regulatory Program** |  |  |  |  |  |  |  |
| **Any** |  |  |  |  |  |  |  |
| 1995-1997 (n=32) | 75.0<br>(56.6-88.5) | 50.0<br>(31.9-68.1) | 31.3<br>(16.1-50.0) | 87.5<br>(71.0-96.5) | 12.5<br>(3.5-29.0) | 34.4<br>(18.6-53.2) | 65.6<br>(46.8-81.4) |
| 2005-2007 (n=36) | 52.8<br>(35.5-69.6) | 36.1<br>(20.8-53.8) | 25.0<br>(12.1-42.2) | 77.8<br>(60.8-89.9) | 22.2<br>(10.1-39.2) | 47.2<br>(30.4-64.5) | 52.8<br>(35.5-69.6) |
| 2015-2017 (n=63) | 38.1<br>(26.1-51.2) | 50.8<br>(37.9-63.6) | 36.5<br>(24.7-49.6) | 73.0<br>(60.3-83.4) | 27.0<br>(16.6-39.7) | 44.4<br>(31.9-57.5) | 55.6<br>(42.5-68.1) |
| 3-way p | P<0.001* | P=0.74 | P=0.48 | P=0.11 |  | P=0.42 |  |
| 2-way p** | P<0.001* | P=0.95 | P=0.62 | P=0.11 |  | P=0.35 |  |
| **None** |  |  |  |  |  |  |  |
| 1995-1997 (n=92) | 82.7<br>(73.3-89.7) | 17.4<br>(10.3-26.7) | 14.1<br>(7.7-23.0) | 98.9<br>(94.1-100.0) | 1.1<br>(0.0-5.9) | 69.6<br>(59.1-78.7) | 30.4<br>(21.3-40.9) |
| 2005-2007 (n=27) | 70.4<br>(49.8-86.2) | 33.3<br>(16.5-54.0) | 11.1<br>(2.4-29.2) | 100.0<br>(87.2-100.0) | 0.0<br>(0.0-12.8) | 51.9<br>(31.9-71.3) | 48.1<br>(28.7-68.1) |
| 2015-2017 (n=43) | 74.4<br>(58.8-86.5) | 39.5<br>(25.0-55.6) | 34.9<br>(21.0-50.9) | 97.7<br>(87.7-99.9) | 2.3<br>(0.0-12.3) | 69.8<br>(53.9-82.8) | 30.2<br>(17.2-46.1) |
| 3-way p | P=0.22 | P=0.004* | P=0.008* | P=0.62 |  | P=0.80 |  |
| 2-way p** | P=0.27 | P=0.006* | P=0.006* | P=0.59 |  | P=0.98 |  |
| **Orphan** |  |  |  |  |  |  |  |
| **Yes** |  |  |  |  |  |  |  |
| 1995-1997 (n=11) | 72.7<br>(39.0-94.0) | 54.5<br>(23.4-83.3) | 36.4<br>(10.9-69.2) | 81.8<br>(48.2-97.7) | 18.2<br>(2.3-51.8) | 81.8<br>(48.2-97.7) | 18.2<br>(2.3-51.8) |
| 2005-2007 (n=17) | 23.5<br>(6.8-49.9) | 52.9<br>(27.8-77.0) | 41.2<br>(18.4-67.1) | 58.8<br>(32.9-81.6) | 41.2<br>(18.4-67.1) | 47.1<br>(23.0-72.2) | 52.9<br>(27.8-77.0) |
| 2015-2017 (n=38) | 23.7<br>(11.4-40.2) | 63.2<br>(46.0-78.2) | 44.7<br>(28.6-61.7) | 65.8<br>(48.6-80.4) | 34.2<br>(19.6-51.4) | 36.8<br>(21.8-54.0) | 63.2<br>(46.0-78.2) |
| 3-way p | P=0.008* | P=0.50 | P=0.61 | P=0.49 |  | P=0.01 |  |
| 2-way p** | P=0.003* | P=0.62 | P=0.63 | P=0.32 |  | P=0.10 |  |
| **No** |  |  |  |  |  |  |  |
| 1995-1997 (n=113) | 81.4<br>(73.0-88.1) | 23.0<br>(15.6-31.9) | 16.8<br>(10.4-25.0) | 97.3<br>(92.4-99.4) | 2.7<br>(0.6-7.6) | 58.4<br>(48.8-67.6) | 41.6<br>(32.4-51.2) |
| 2005-2007 (n=46) | 73.9<br>(58.9-85.7) | 28.3<br>(16.0-43.5) | 10.9<br>(3.6-23.6) | 97.8<br>(88.5-99.9) | 2.2<br>(0.0-11.5) | 50.0<br>(34.9-65.1) | 50.0<br>(34.9-65.1) |
| 2015-2017<br>(n=68) | 69.1<br>(56.7-79.8) | 36.8<br>(25.4-49.3) | 30.9<br>(20.2-43.3) | 92.6<br>(83.7-97.6) | 7.4<br>(2.4-16.3) | 64.7<br>(52.2-75.9) | 35.3<br>(24.1-47.8) |
| 3-way p | P=0.06 | P=0.05 | P=0.04 | P=0.14 |  | P=0.50 |  |
| 2-way p** | P=0.06 | P=0.05 | P=0.03 | P=0.14 |  | P=0.40 |  |

\* indicates $p < 0.025$ (after Bonferroni adjustment for 2 comparisons)
\*\* 2-way p-value for differences between 1995-1997 and 2015-2017 time periods.

**Table 5.**
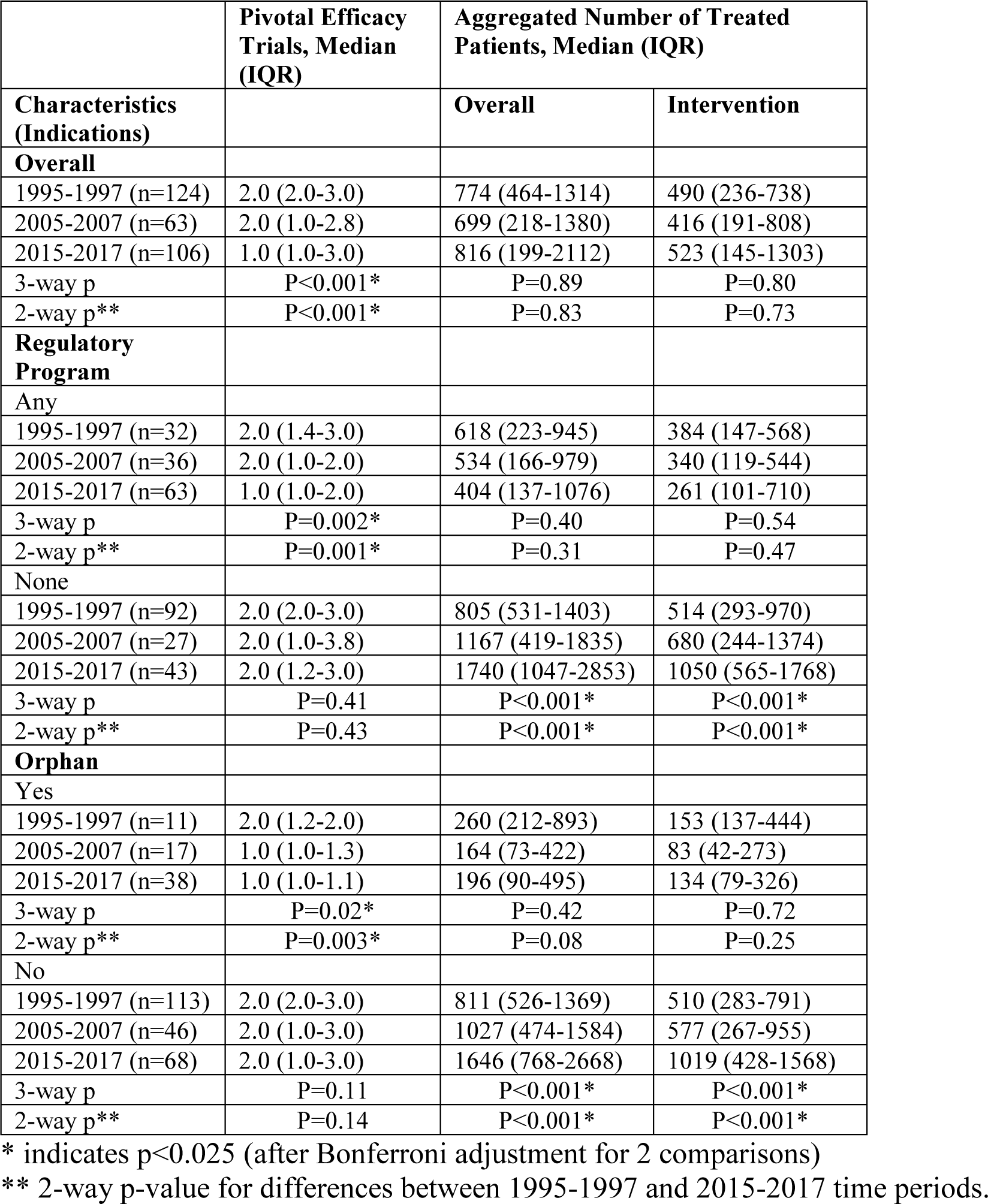
Number of Pivotal Efficacy Trials and Number of Treated Patients in Aggregated Pivotal Trials Supporting U.S. Food and Drug Administration Indication Approvals of New Drugs and Biologics in 1995-1997, 2005-2007, and 2015-2017, Stratified by Drug/Indication Characteristics.

### Aggregated Pivotal Trial Features Supporting Indication Approvals by Use of Special Regulatory Programs

The proportion of new drug or biologic approvals using any special regulatory program (Priority Review, Accelerated Approval, Fast Track, or Breakthrough Designation) increased over time, with 37 (34.6%) in 1995-1997, 33 (57.9%) in 2005-2007, and 70 (64.2%) in 2015-2017 (p<0.001). Among indication approvals using any special regulatory program, the proportion supported by at least 2 pivotal trials decreased over time (75.0% in 1995-1997, 52.8% in 2005-2007, 38.1% in 2015-2017; p<0.001) **(Table 4)**. There were not statistically significant changes in the proportion supported only by single-arm trials (p=0.11), only by trials using surrogate endpoints (p=0.42), by at least one trial of 6 months’ duration (p=0.74), or in the median aggregated number of treated patients (p=0.40) **(Tables 4-5)**.

Among indication approvals not using any special regulatory program, there were not statistically significant changes in the proportion supported by at least 2 pivotal trials (p=0.22), only by single-arm trials (p=0.62), or by trials using surrogate endpoints (p=0.80) **(Table 4)**. However, the median aggregated number of treated patients increased over time (805 [IQR 531-1403] in 1995-1997, 1167 [IQR 419-1835] in 2005-2007, 1740 [IQR 1047-2853] in 2015-2017; p<0.001), as did the proportion supported by at least one trial of 6 months’ duration (17.4% in 1995-1997, 33.3% in 2005-2007, 39.5% in 2015-2017; p=0.004) **(Tables 4-5)**.

### Aggregated Pivotal Trial Features Supporting Indication Approvals by Orphan Designation Status

The proportion of indication approvals receiving orphan designations increased over time, with 20 (12.7%) in 1995-1997, 17 (26.6%) in 2005-2007, and 45 (38.1%) in 2015-2017 (p<0.001). Trends in the features of aggregated pivotal trials supporting indications over time differed when stratified by orphan designation status. Among orphan indications, the proportion of indication approvals supported by at least 2 pivotal trials decreased over time (72.7% 1995-1997, 23.5% in 2005-2007, 23.7% in 2015-2017; p=0.008) **(Table 4)**. The proportion of orphan indications supported only by single-arm trials did not change (p=0.49), while those supported only by trials using surrogate endpoints (18.2% in 1995-1997, 52.9% in 2005-2007, 63.2% in 2015-2017; p=0.01) increased over time. The median aggregated number of treated patients for orphan indications did not differ over time (p=0.42), nor did the proportion of indications with at least one trial of 6 months’ duration (p=0.50) **(Tables 4-5)**.

In contrast, the proportion of non-orphan indications supported by at least 2 pivotal trials did not change over time (p=0.06), nor did those supported only by single-arm trials (p=0.14), those supported only by trials using surrogate endpoints (p=0.50), or those supported by at least one trial of 6 months’ duration (p=0.05). Meanwhile, the median aggregated number of treated patients increased (811 [IQR 526-1369] in 1995-1997, 1027 [IQR 474-1584] in 2005-2007, 1646 [IQR 768-2668] in 2015-2017; p<0.001).

### Supplementary and Sensitivity Analyses

Supplementary analyses for individual pivotal trial and aggregated indication approval characteristics were stratified by product type and therapeutic area, and expected length of treatment **(Tables S6-9)**. Sensitivity analyses were conducted examining trial duration stratified by expected length of treatment **(Tables S10-11)**. Trends in aggregated indication approval characteristics over time were consistent when stratified by Priority Review and Accelerated Approval considered separately (**Tables S12-13)** when compared to special regulatory programs considered as a whole, as were trends when considering orphan designation as a special regulatory program **(Tables S14-15)**.

## DISCUSSION

We reviewed all new drugs and biologics approved by the FDA in 1995-1997, 2005-2007, and 2015-2017 and found differences over time in the number and quality of the pivotal trials supporting their approval. The aggregated evidence supporting indication approvals has become less rigorous in some ways, with the proportion of approvals supported by the commonly-understood standard of “at least 2 pivotal trials” declining from 81% to 53% and the proportion of approvals supported by at least one trial using a comparator declining from 96% to 83%. Meanwhile, it has become more rigorous in other ways, with the proportion of indications supported by at least one trial of 6 months’ duration increasing from 26% to 46%. These findings have implications for patients and clinicians making decisions as to whether to use products newly available on the market, as well as reinforcing the importance of FDA’s strategy for requiring ongoing evaluation of therapeutic safety and efficacy after approval.

### Policy Considerations

A key question is whether these overall trends are driven by changes in the frequency with which approvals use special regulatory programs or orphan designation, or if there has been an evolution in the standards needed to secure approval over time. Our findings suggest both explanations may play a role. We found a significant increase in the proportion of new indications approved using any special regulatory program or orphan designation, in line with previous reports [7]. These changes are likely reflected in the overall trends regarding the evidence supporting approvals, given that the use of special regulatory programs and orphan designation are both associated with more flexible standards for approval [5,18]. Importantly, however, we also found changes in the aggregated evidence over time even when accounting for use of these programs, suggesting potential involvement of factors beyond compositional effects. There was a decrease in the number of pivotal trials supporting an indication approval only among therapeutics using a special regulatory program, while therapeutics not using a special regulatory program showed increases in the aggregated number of treated patients and the proportion of indications supported by a trial of at least 6 months’ duration. These trends were consistent when examining individual associations with Priority Review, Accelerated Approval, and Orphan Designation, which were used in all three time periods studied.

These divergent patterns in evidentiary requirements highlight the trade-offs inherent to special regulatory programs and orphan status. These programs are intended to encourage development in areas where there are clinically unmet needs, and accordingly facilitate approvals based on fewer trials, shorter trials, or trials using surrogate markers of disease as endpoints, thus requiring less time to ascertain an effect and enabling products to reach the market sooner. The proliferation of the use of these programs over the past three decades, with their flexibility in pre-market standards, reinforces the importance of a life-cycle approach to evaluating drug efficacy and safety in today’s regulatory environment [19]. Proponents of this approach embrace the use of post-marketing studies [20], pragmatic trials, and real-world evidence (RWE) to ensure continued evidence development for new medical products. In part spurred by the 21st Century Cures Act, the FDA has particularly embraced the use of RWE – defined expansively to include clinical evidence obtained from a variety of settings, including electronic health records, insurance claims, registries, and personal devices [21] – as a new frontier of regulatory science. In the past few years, the FDA has furthered the integration of RWE into its medical product evaluation process, issuing guidance on the use of RWE in decision-making about drugs, biologics, and devices [22,23]. Continued development of life-cycle evaluation methods, including enhanced requirements that ensure that studies are undertaken and reported, may strengthen efforts to generate clinical evidence on the safety and efficacy of drugs and biologics after approval, to inform patients, clinicians, and regulators.

### Limitations

Our study had several limitations. First, our study included only three-year samples of approvals in each time period and cannot capture the full range of therapeutic agents and variations in approval trends across entire decades. Second, given our sample, we could not fully adjust for interactions between all combinations of drug attributes and special regulatory programs and orphan status. For instance, it was not uncommon for a therapeutic approval to use multiple special regulatory programs, and we could not fully account for the possibility that a single regulatory program or attribute had disproportionate influence on the associations observed. However, the consistency of our findings regarding special regulatory programs across multiple individual special regulatory programs, as well as orphan designation, suggests that we were able to capture major trends. Lastly, we included only clinical trials identified as pivotal trials in our study. Other non-pivotal studies and data, such as observational studies or previous marketing experience from other countries, may contribute to FDA reviewers’ holistic evaluations of drugs under consideration in ways that cannot be captured by our approach.

## CONCLUSION

The quality of clinical trial evidence used to support new drug and biologic approvals has changed over the past three decades, requiring on average fewer pivotal trials with less robust comparators, but with longer durations. This change has implications for physicians and patients as they consider using newly-approved drugs, as well as for regulators, as it reinforces the importance of FDA’s strategy for requiring ongoing evaluation of therapeutic safety and efficacy after approval.

## Data Availability

Data will be posted publicly upon publication.

## ACKNOWLEDGEMENTS

### Contributors

ADZ, JSR conceived and designed this study. ADZ, JP, NSD acquired the data. ADZ conducted the statistical analysis and drafted the manuscript. All authors participated in the interpretation of the data and critically revised the manuscript for important intellectual content. ADZ and JSR had full access to all the data in the study and take responsibility for the integrity of the data and the accuracy of the data analysis. JSR provided supervision. ADZ and JSR are guarantors.

### Funding

This project was not supported by any external grants or funds.

### Competing interests

All authors have completed the <u>Unified Competing Interest form</u> (available on request from the corresponding author) and declare: In the past 36 months, Ms. Zhang received research support as a scholar in the Yale-Mayo Clinic FDA CERSI (U01FD005938) and from the Laura and John Arnold Foundation. In the past 36 months, Mr. Puthumana received a student research grant provided by the Yale School of Medicine Office of Student Research (National Institutes of Health training grant T35DK104689). Dr. Downing is currently employed by Bain Capital Life Sciences, a venture company. His participation in this work took place during his postgraduate training in internal medicine at the Brigham & Women’s Hospital. In the past 36 months, Dr. Krumholz was a recipient of a research grant, through Yale, from Medtronic and the U.S. Food and Drug Administration to develop methods for post-market surveillance of medical devices; was a recipient of a research grant with Medtronic and Johnson & Johnson, through Yale, to develop methods of clinical trial data sharing; was a recipient of a research agreement, through Yale, from the Shenzhen Center for Health Information for work to advance intelligent disease prevention and health promotion; collaborates with the National Center for Cardiovascular Diseases in Beijing; received payment from the Arnold & Porter Law Firm for work related to the Sanofi clopidogrel litigation and from the Ben C. Martin Law Firm for work related to the Cook IVC filter litigation; chairs a Cardiac Scientific Advisory Board for UnitedHealth; is a participant/participant representative of the IBM Watson Health Life Sciences Board; is a member of the Advisory Board for Element Science, the Advisory Board for Facebook, and the Physician Advisory Board for Aetna; and is the founder of Hugo, a personal health information platform. In the past 36 months, Dr. Shah has received research support through Mayo Clinic from the Food and Drug Administration to establish Yale-Mayo Clinic Center for Excellence in Regulatory Science and Innovation (CERSI) program (U01FD005938); the Centers of Medicare and Medicaid Innovation under the Transforming Clinical Practice Initiative (TCPI); the Agency for Healthcare Research and Quality (R01HS025164; R01HS025402; R03HS025517); the National Heart, Lung and Blood Institute of the National Institutes of Health (NIH) (R56HL130496; R01HL131535); the National Science Foundation; and the Patient Centered Outcomes Research Institute (PCORI) to develop a Clinical Data Research Network (LHSNet). In the past 36 months, Dr. Ross has received research support through Yale University from Johnson and Johnson to develop methods of clinical trial data sharing, from Medtronic, Inc. and the Food and Drug Administration (FDA) to develop methods for postmarket surveillance of medical devices (U01FD004585), from the Food and Drug Administration to establish Yale-Mayo Clinic Center for Excellence in Regulatory Science and Innovation (CERSI) program (U01FD005938), from the Blue Cross Blue Shield Association to better understand medical technology evaluation, from the Centers of Medicare and Medicaid Services (CMS) to develop and maintain performance measures that are used for public reporting (HHSM-500-2013-13018I), from the Agency for Healthcare Research and Quality (R01HS022882), from the National Heart, Lung and Blood Institute of the National Institutes of Health (NIH) (R01HS025164), and from the Laura and John Arnold Foundation to establish the Good Pharma Scorecard at Bioethics International and to establish the Collaboration for Research Integrity and Transparency (CRIT) at Yale.

### Patient consent

Not required.

### Ethical approval

Not required.

### Data sharing

Data can be shared upon request.

### Transparency

The manuscript guarantors (ADZ and JSR) affirm that this manuscript is an honest, accurate, and transparent account of the study being reported; that no important aspects of the study have been omitted; and that any discrepancies from the study as planned (and, if relevant, registered) have been explained.

### License

The Corresponding Author has the right to grant on behalf of all authors and does grant on behalf of all authors, an exclusive licence (or non-exclusive for government employees) on a worldwide basis to the BMJ Publishing Group Ltd to permit this article (if accepted) to be published in BMJ editions and any other BMJPGL products and sublicences such use and exploit all subsidiary rights, as set out in our licence.

## REFERENCES

1 Drug Amendments of 1962. 1962.

2 US FDA. Guideline for the Format and Content of the Clinical and Statistical Sections of an Application. 1988. https://www.fda.gov/downloads/Drugs/GuidanceComplianceRegulatoryInformation/Guidances/UCM071665.pdf (accessed 6 Dec 2018).

3 US FDA. Providing Clinical Evidence of Effectiveness for Human Drug and Biological Products: Guidance for Industry. 1998. https://www.fda.gov/downloads/drugs/guidancecomplianceregulatoryinformation/guidances/ucm072008.pdf (accessed 14 Dec 2018).

4 Puthumana J, Wallach JD, Ross JS. Clinical Trial Evidence Supporting FDA Approval of Drugs Granted Breakthrough Therapy Designation. JAMA 2018;320:301–3. doi:10.1001/jama.2018.7619

5 Downing NS, Aminawung JA, Shah ND, et al. Clinical Trial Evidence Supporting FDA Approval of Novel Therapeutic Agents, 2005-2012. JAMA 2014;311:368–377. doi:10.1001/jama.2013.282034

6 Pregelj L, Hwang TJ, Hine DC, et al. Precision Medicines Have Faster Approvals Based On Fewer And Smaller Trials Than Other Medicines. Health Aff (Millwood) 2018;37:724–31. doi:10.1377/hlthaff.2017.1580

7 Kesselheim AS, Wang B, Franklin JM, et al. Trends in utilization of FDA expedited drug development and approval programs, 1987-2014: cohort study. BMJ 2015;351:h4633. doi:10.1136/bmj.h4633

8 US FDA. Drugs@FDA: FDA Approved Drug Products. 2018.https://www.accessdata.fda.gov/scripts/cder/daf/ (Accessed 31 Oct 2018).

9 US FDA CDER. NDA and BLA Approval Reports - Priority NDA and BLA Approvals. 2018.https://www.fda.gov/Drugs/DevelopmentApprovalProcess/HowDrugsareDevelopedandApproved/DrugandBiologicApprovalReports/NDAandBLAApprovalReports/ucm2007012.htm (accessed 10 Dec 2018).

10 US FDA CDER. NDA and BLA Approval Reports - Accelerated Approvals. 2018.https://www.fda.gov/Drugs/DevelopmentApprovalProcess/HowDrugsareDevelopedandApproved/DrugandBiologicApprovalReports/NDAandBLAApprovalReports/ucm373430.htm (accessed 10 Dec 2018).

11 US FDA CDER. NDA and BLA Approval Reports - Fast Track Approvals for Drugs, 1998-2006. 2018.https://www.fda.gov/Drugs/DevelopmentApprovalProcess/HowDrugsareDevelopedandApproved/DrugandBiologicApprovalReports/NDAandBLAApprovalReports/ucm2007016.htm (accessed 10 Dec 2018).

12 US FDA CDER. NDA and BLA Approval Reports - Breakthrough Therapy Approvals. 2018.https://www.fda.gov/Drugs/DevelopmentApprovalProcess/HowDrugsareDevelopedandApproved/DrugandBiologicApprovalReports/NDAandBLAApprovalReports/ucm373418.htm (accessed 10 Dec 2018).

13 US FDA. Guidance for Industry: Expedited Programs for Serious Conditions – Drugs and Biologics. 2014.

14 Food and Drug Administration Safety and Innovation Act. 2012. https://www.gpo.gov/fdsys/pkg/PLAW-112publ144/pdf/PLAW-112publ144.pdf (accessed 24 Oct 2018).

15 WHO. World Health Organization (WHO) Collaborating Centre for Drug Statistics Methodology. 2018.https://www.whocc.no/atc_ddd_index/ (Accessed 31 Oct 2018).

16 US FDA. Orphan Drug Designations and Approvals. 2018.https://www.accessdata.fda.gov/scripts/opdlisting/oopd/index.cfm (Accessed 31 Oct 2018).

17 Pease AM, Krumholz HM, Downing NS, et al. Postapproval Studies of Drugs Initially Approved by the FDA on the Basis of Limited Evidence: Systematic Review. BMJ 2017;357:j1680. doi:10.1136/bmj.j1680

18 Kesselheim AS, Myers JA, Avorn J. Characteristics of Clinical Trials to Support Approval of Orphan vs. Nonorphan Drugs for Cancer. JAMA 2011;305:2320–2326. doi:10.1001/jama.2011.769

19 Institute of Medicine (US) Forum on Drug Discovery and Devleopment. Integrating Pre- and Postmarket Review. National Academies Press (US) 2007. https://www.ncbi.nlm.nih.gov/books/NBK52931/ (accessed 4 Jun 2019).

20 Wallach JD, Ross JS, Naci H. The US Food and Drug Administration’s expedited approval programs: Evidentiary standards, regulatory trade-offs, and potential improvements. Clin Trials 2018;15:219–29. doi:10.1177/1740774518770648

21 Sherman RE, Anderson SA, Dal Pan GJ, et al. Real-World Evidence - What Is It and What Can It Tell Us? NEJM 2016;375:2293–2297. doi:10.1056/NEJMsb0903885

22 US FDA. Use of Real-World Evidence to Support Regulatory Decision-Making for Medical Devices: Draft Guidance. 2016.

23 US FDA. Submitting Documents Using Real-World Data and Real-World Evidence to FDA for Drugs and Biologics Guidance for Industry. 2019.

